# Clinical Performance of SARS-CoV-2 Rapid Antigen Tests: A Systematic Review and Meta-Analysis

**DOI:** 10.1101/2023.09.15.23295560

**Authors:** Nina Germic, Johannes Hayer, Qian Feng

**Affiliations:** Roche Diagnostics International Ltd, Rotkreuz, Switzerland; Roche Diagnostics GmbH, Mannheim, Germany

**Keywords:** SARS-CoV-2, COVID-19, clinical performance, rapid antigen test, lateral flow assay, point-of-care, in vitro diagnostics

## Abstract

**Objectives:** We conducted a meta-analysis of RAT diagnostic accuracy for SARS-CoV-2 infections, and further evaluated test sensitivity versus the presence of symptoms, days post symptom onset (DPSO), sample viral load, and sample type (i.e. direct swabs versus specimens stored in transport media).

**Methods:** Three databases were searched systematically for performance evaluations of the Roche-distributed SDB SARS-CoV-2 Rapid Antigen Test (Roche/SDB RAT) through March 2022. If the Roche/SDB RAT was compared with any of 9 commonly available antigen tests, data from these tests were also included.

**Results:** Overall sensitivity of RATs among different manufacturers and study cohorts varied between 36.0% (95% CI: 24.0-50.1) and 79.4% (95% CI: 64.8-89.0). Roche/SDB RATs demonstrated a competitive performance with a pooled (including off-label use) sensitivity of 70.0%, and nearly 100% specificity in included studies. The Roche/SDB RATs exhibited reliable sensitivity in patients with a relatively high viral load (96.6% [95% CI: 95.2-98.2] for Ct≤25). Roche/SDB RATs were more sensitive in symptomatic patients within the first 7 DPSO (85.5% [95% CI: 81.2-88.4]), and when used to test direct swabs (74.4% [95% CI: 69.7-80.3]).

**Conclusion:** RATs show reliable performance in clinical settings and should be considered when rapid diagnosis of SARS-CoV-2 infection is critical.

**HIGHLIGHTS:**

- Meta-analysis of 86 studies of SARS-CoV-2 rapid antigen test (RAT) performance
- RAT performance supports near-patient testing for early COVID-19 diagnosis
- RAT specificity is high and sensitivity is reliable in those with high viral load
- RAT sensitivity in symptomatic patients is higher than in asymptomatic individuals
- RAT sensitivity is higher for direct swabs compared to swabs in transport media

## INTRODUCTION

Coronavirus disease 2019 (COVID-19) is caused by infection with severe acute respiratory syndrome coronavirus 2 (SARS-CoV-2)[1] and as of August 2023, there have been more than 769 million confirmed cases of COVID-19, including 6.9 million deaths reported to WHO.[2] The clinical spectrum of SARS-CoV-2 infection is wide, ranging from an asymptomatic infection to severe pneumonia with acute respiratory distress syndrome and death.[3–5] Timely diagnosis can contribute to clinical management and outbreak control. Diagnostic testing involves detecting viral biomarkers, or detecting the human immune response to the virus.[6] Commonly used diagnostic methods for SARS-CoV-2 include Real-Time Polymerase Chain Reaction (RT-PCR), which has become the gold standard, viral culture, serology assays, and antigen-detection tests, both at Point-of-Care and in laboratories.[7] RT-PCR enables accurate diagnosis of SARS-CoV-2 infection with technical turn-around times ranging from less than an hour to 24 hours.[8]

Rapid antigen tests (RATs) produce results quickly with a turnaround time of 15-30 minutes.[9] Further, RATs are convenient to use, widely available to the general population, and less expensive than NAATs.[9] However, they are typically less sensitive than RT-PCR and other NAATs. Varying clinical performance has been reported for SARS-CoV-2 RATs from different manufacturers and among diverse patient populations on a global scale. A previous meta-analysis reported a pooled RAT sensitivity and specificity of 71.2% and 98.9% respectively, although sensitivity increased to 76.3% when analysis was confined to studies that followed the manufacturers’ instructions.[10] A Cochrane review also summarized results from various studies that examined or analyzed the diagnostic accuracy of rapid antigen tests.[11] When tests were used according to manufacturer instructions, average sensitivities by brand ranged from 34.3% to 91.3% in symptomatic participants for 20 tests with eligible data, while sensitivity ranged from 28.6% to 77.8% for 12 tests studied in asymptomatic participants. Corman et al. showed that the Abbott Panbio™ COVID-19 Ag Rapid Test, the Healgen^®^ Coronavirus Ag Rapid Test Cassette (Swab), the R-Biopharm RIDA^®^ QUICK SARS-CoV-2 Antigen Test, and the Roche/SD Biosensor SARS-CoV-2 Rapid Antigen Test detect as few as 4·4 plaque-forming units (PFU) of virus (88 PFU/mL).[12]

The SARS-CoV-2 Rapid Antigen Test manufactured by SD Biosensor and distributed by Roche Diagnostics (equivalent to the STANDARD Q COVID-19 Ag Test; hereafter “Roche/SDB RAT”)^1^ was broadly used internationally during the pandemic, both in professional point-of-care settings and as a self-test. To evaluate the diagnostic accuracy of the Roche/SDB RAT, we performed an unbiased literature search including studies across 36 different countries.[12] In this meta-analysis, we analyzed the overall performance of the Roche/SDB RAT, followed by sub-analyses according to the presence of symptoms, days post symptom onset (DPSO), cycle threshold (Ct) ranges and sample types (direct swabs vs samples stored in transport media). If the Roche/SDB RAT was compared with one of 9 other widely available antigen tests (Abbott, Acon, BD, Biosynex, Boson, Laihe, MP BIO, Siemens, Quidel), data from these tests were included as well.

Our meta-analysis of 86 studies shows that RAT performance supports near-patient testing for early COVID-19 diagnosis, with reliable sensitivity in those with relatively high viral load. In general, RATs correctly diagnosed the bulk of SARS-CoV-2-infected persons within the first week of symptom onset.

## OBJECTIVES AND STUDY DESIGN

### Search Strategy

This meta-analysis was performed according to the Preferred Reporting Items for Systematic Reviews and Meta-Analyses (PRISMA) guidelines.[13] SURUS, a custom-built natural language processing (NLP) Engine (Medstone Science B.V.) was used to conduct query-based searches for relevant papers from January 2020 through March 2022. The databases MEDLINE, and preprint servers MedRixv and BioRxiv were initially searched for clinical performance studies of a commercial SARS-CoV-2 RAT with the following search strings: (“SARS-CoV-2” OR “Rapid antigen test” OR “point of care” or “lateral flow assay” or “Roche/SD Biosensor/Standard Q”) AND (“nasopharyngeal” OR “nasal” OR “oro-nasopharyngeal” OR “oropharyngeal” OR “viral culture”) AND (“Sensitivity” OR “Specificity” OR “Accuracy” OR “PPA” OR “NPA” OR “PPV” OR “NPV” OR “LOD” OR “TCID”). Only papers that evaluated the Roche/SDB RAT performance with any of the specified parameters were considered if results were reported at the manufacturer level. If the Roche/SDB RAT was compared with one of the 9 other antigen tests of interest (Abbott, Acon, BD, Biosynex, Boson, Laihe, MP BIO, Siemens, Quidel), the data from these tests were included as well.

In addition to the machine-learning approach, a manual search including FIND (The Foundation for Innovative New Diagnostics [FIND], 2020) was conducted.

### Data Extraction

Data extraction from identified studies was carried out by Medstone Science B.V. using the NLP algorithm, and results were manually validated and verified. The following information was extracted: publication characteristics (e.g., publication date, publication type, publication ID, and hyperlink to full text), study design (e.g., study size, location, timelines, sample collection, reference test), characteristics of study participants (e.g., age, presence of symptoms, DPSO at the time of diagnosis), test performance (e.g., sensitivity and specificity) and QUADAS parameters for study quality.[14] We considered 7 quality parameters that are known to affect test performance, including 1) whether the patient sample represented the target population; 2) whether patient selection criteria were clearly described; 3) whether the time interval between the reference test and RAT was appropriately brief; 4) whether RAT execution was described in sufficient detail for test replication; 5) whether reference test execution was described in sufficient detail for test replication; 6) whether blinded interpretation of RAT results occurred; and 7) whether patient withdrawals and sample exclusion were explained.

### Selection criteria

Initially, 135 records were identified. We excluded duplicates, records that were not in the English language or outside the analysis time window, and any publications that did not evaluate the Roche/SDB RAT, thus retaining 124 publications for further analysis. All 124 publications used an RT-PCR test as a comparator.

For qualitative analysis, studies were excluded if:

a) the total number of analyzed samples was <100; or
b) testing was not performed using clinical samples (i.e. contrived or cultured viral material); or
c) the specimen source for the antigen testing was only “saliva”; or
d) time between antigen sampling and comparator sampling was defined as “> 24 hours”.

Ninety-seven records were identified for qualitative analysis.

For quantitative analysis (meta-analysis), studies were excluded if no discernible numbers of true positive (TP) and false negative (FN), or true negative (TN) and false positive (FP) values could be manually extracted from the presented data in the eligible studies. Eighty-six records were identified for quantitative analysis.

### Statistical Analysis

The extracted data were entered into an electronic database and classified according to the RAT that was studied. Information on the overall clinical performance (sensitivity, specificity) and stratified analysis based on DPSO, presence or absence of symptoms at the time of sampling, Ct values of paired RT-PCR results, and use of direct swabs vs specimens stored in transport media was recorded for each eligible study.

As confidence intervals (CIs) reported in the publications were calculated using differing methods, all confidence intervals were recalculated using the exact Clopper– Pearson method for better comparability. Due to the heterogeneity in sub-groups and the small number of studies available for some RATs, we report the differences between tests descriptively rather than statistically. Relative sensitivity and specificity of RATs were calculated in relation to RT-PCR results as the gold standard (based on the number of TP, TN, FP, and FN values extracted from eligible studies).

The meta-analysis of the performance results of RATs against RT-PCR reference methods was performed using the statistical software R (R Foundation for Statistical Computing, 2020).

For overall clinical performance, we undertook the statistical pooling of estimates across all manufacturers namely Abbott, Acon, BD, Biosynex, Boson, Laihe, MP BIO, Roche, Siemens, and Quidel. The metaprop function from the “meta” package[15] was used to calculate the effect size for each individual test and pooled overall in a forest plot (**Figure 1**).

**Figure 1.**
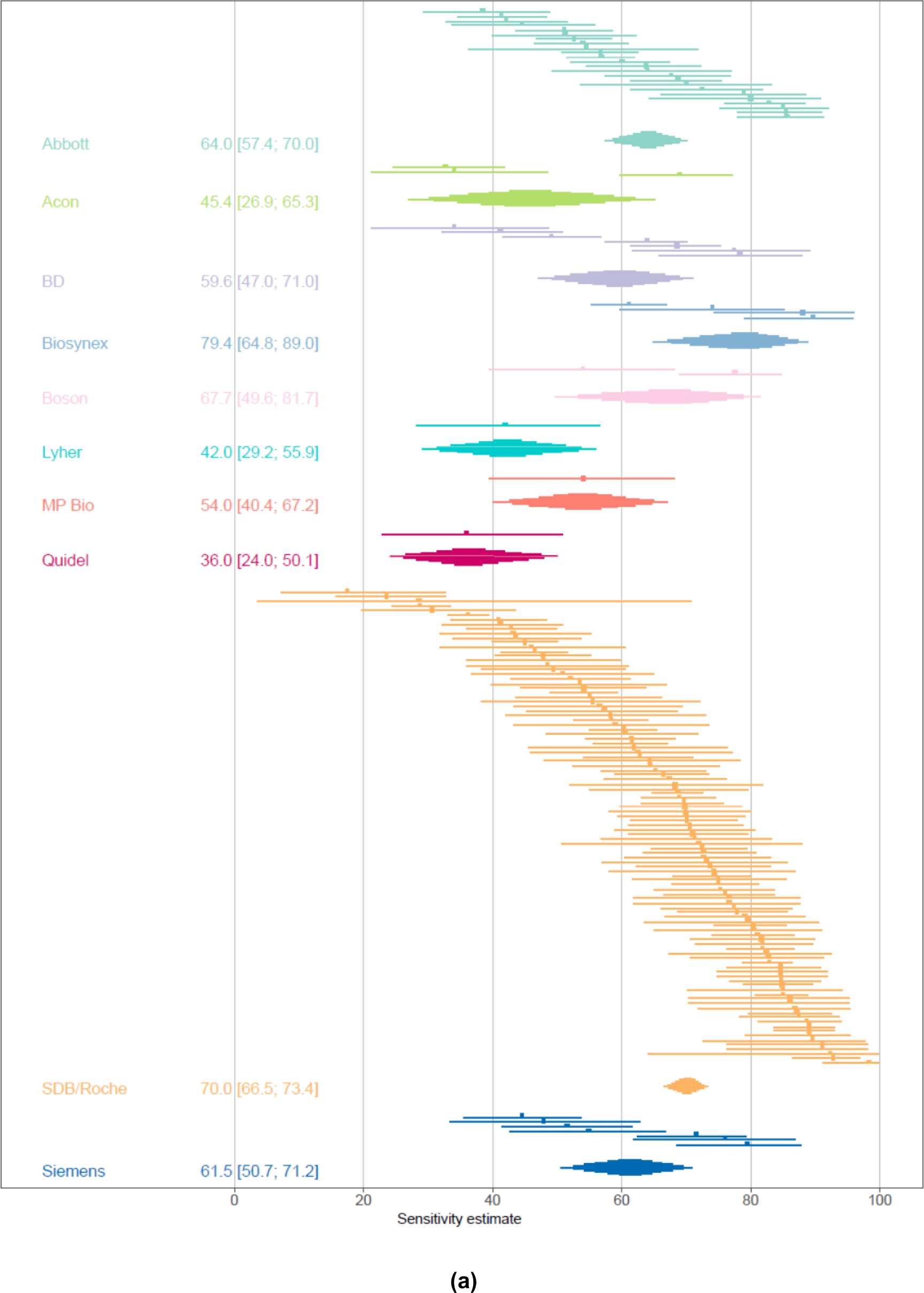
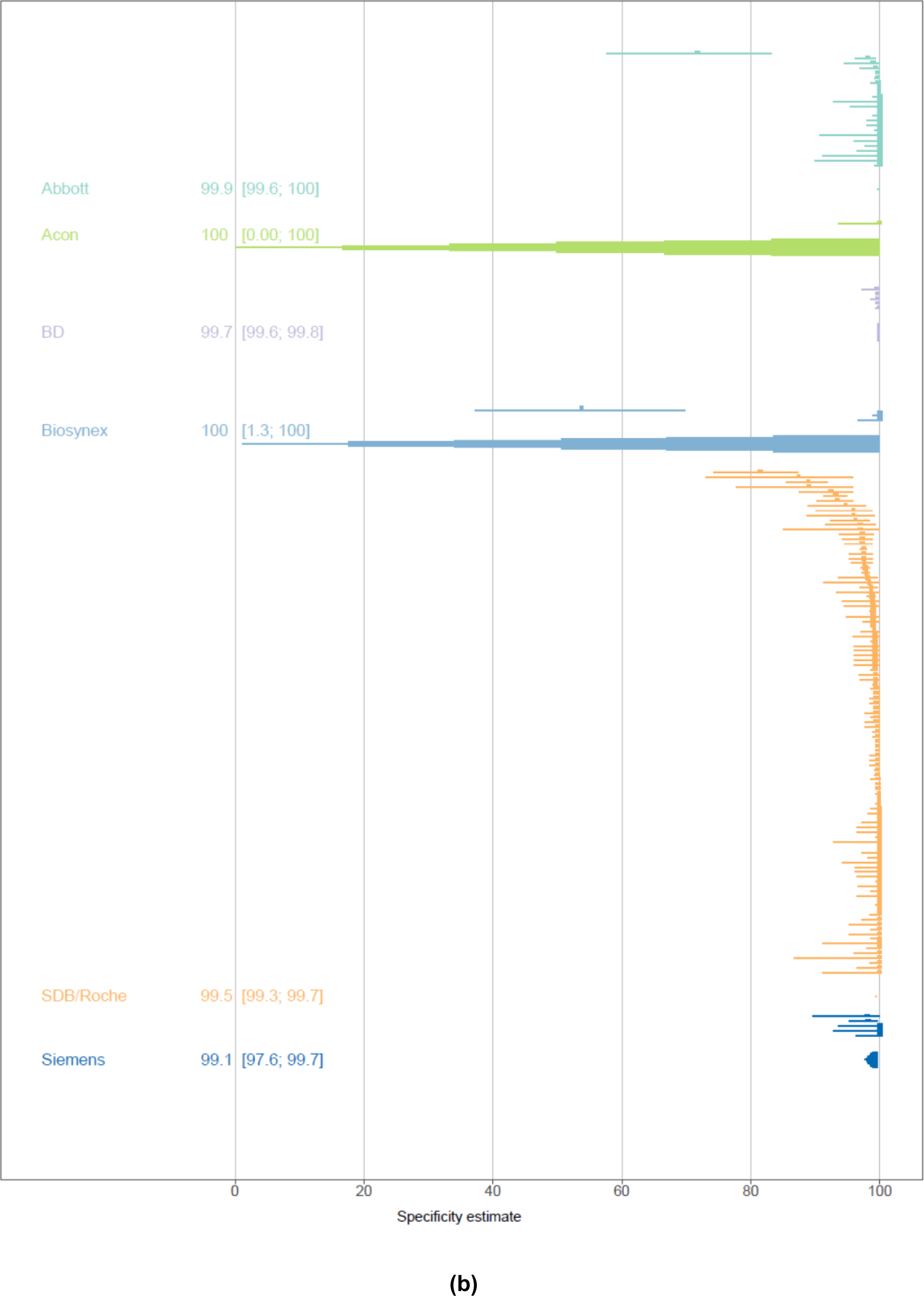
(a) Forest plot of included studies showing overall sensitivity across different RATs. Each row corresponds to one study (if distinct cohorts were presented within one study, the sensitivity is presented per cohort). Sensitivity is marked by a dot and the corresponding 95% confidence interval by a line, with the results ordered by test manufacturer. Additionally, the summary result for each manufacturer is depicted by a diamond at the end of each section. (b) Forest plot of studies evaluating overall specificity across RAT manufacturers.

The stratified analyses discussed above are depicted as forest plots (**Figures 2 and 3**) which show the individual results of each study. Due to the high heterogeneity of the results in the different studies, rather than the mean, the median and a corresponding CI were used in the result descriptions. The CIs were calculated using a Wald interval on the ranks. In case the sample size was not sufficient for the calculations, the minimal and maximal study result per condition (and manufacturer) was used as a proxy for confidence limits.

**Figure 2:**
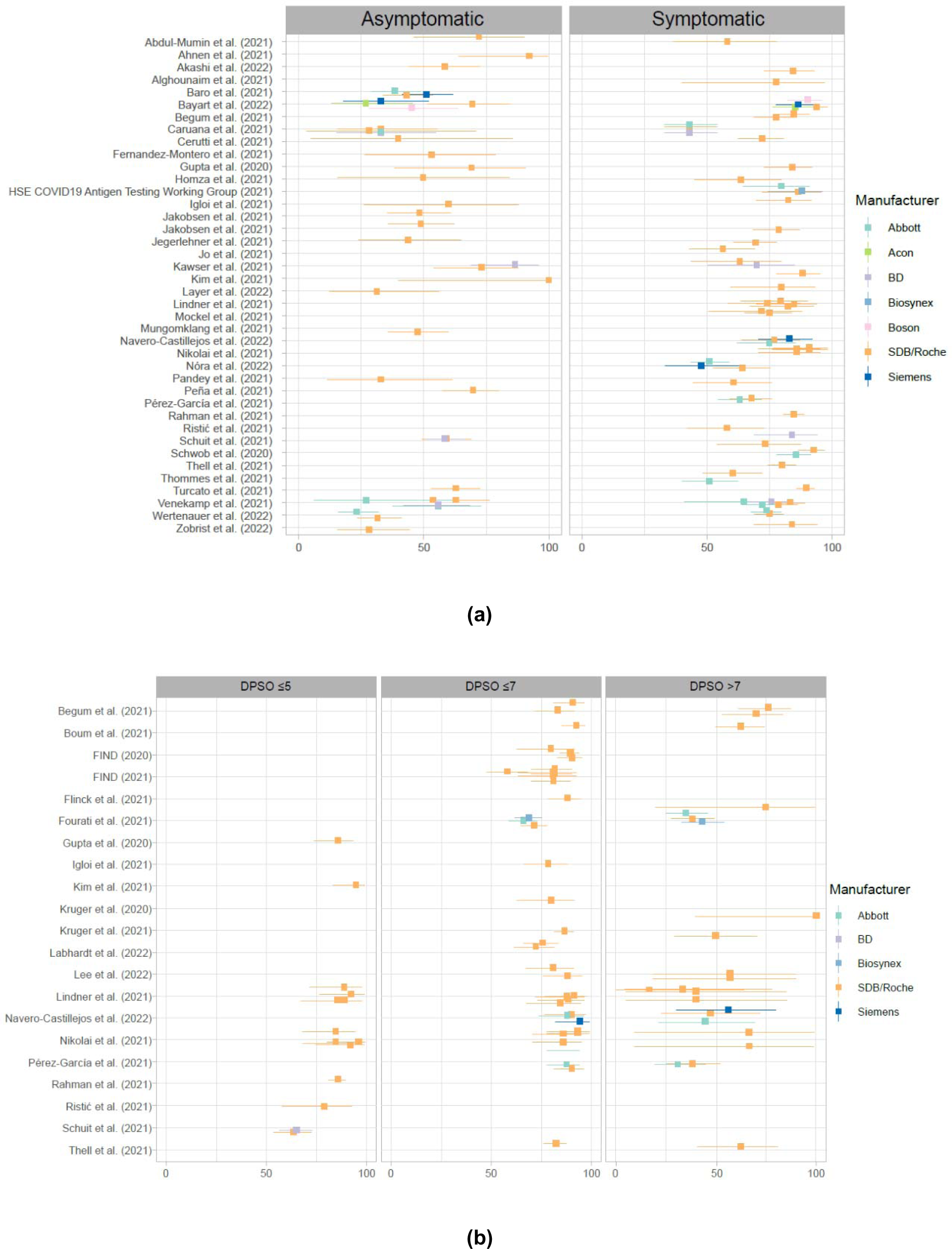
(a) Forest plot of studies showing sensitivity for symptomatic and asymptomatic patients, grouped by RAT manufacturer. (b) Forest plot of studies showing sensitivity grouped by Days Post Symptom Onset (DPSO) threshold (≤5, ≤7 and >7).

**Figure 3:**
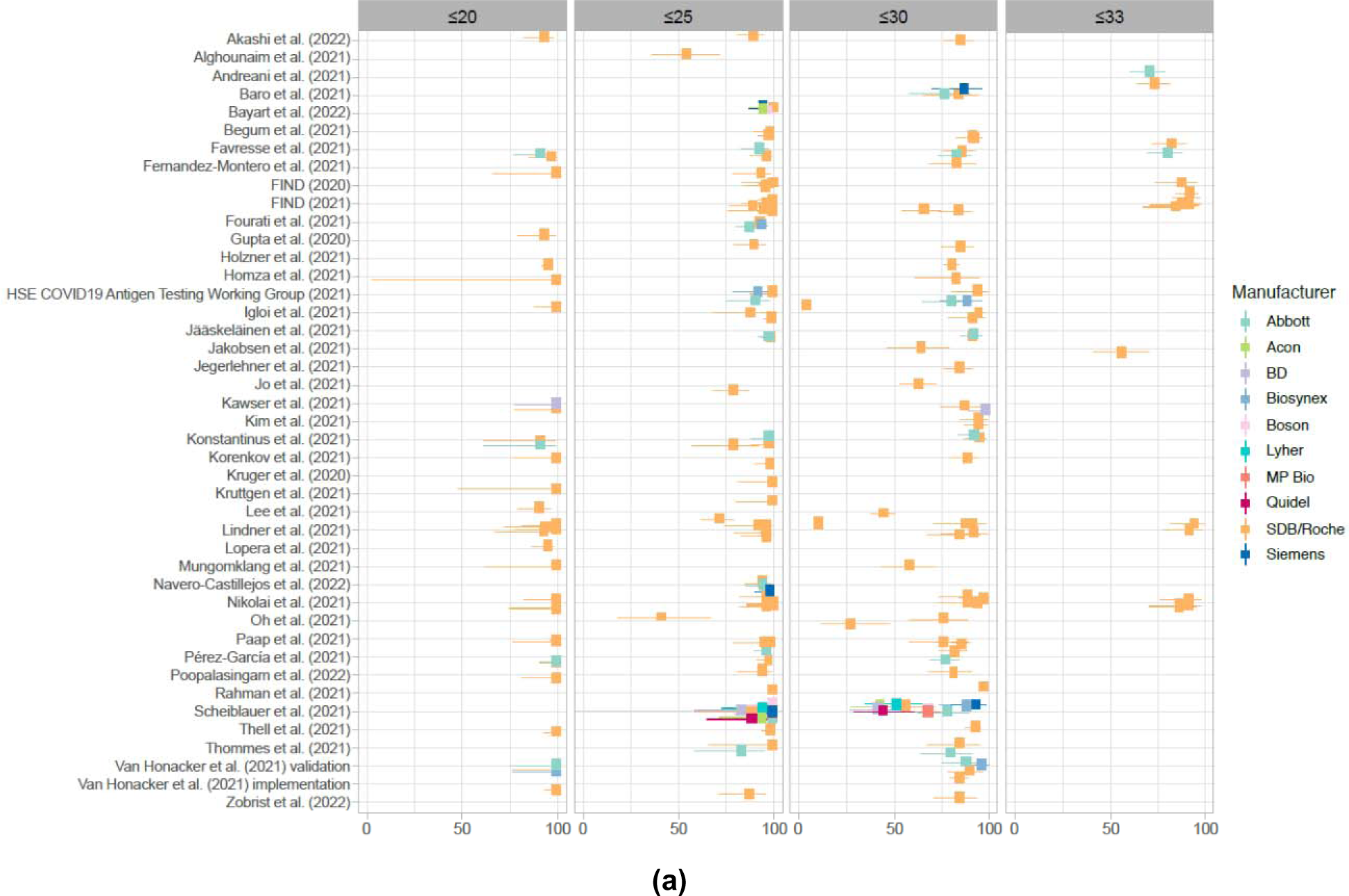
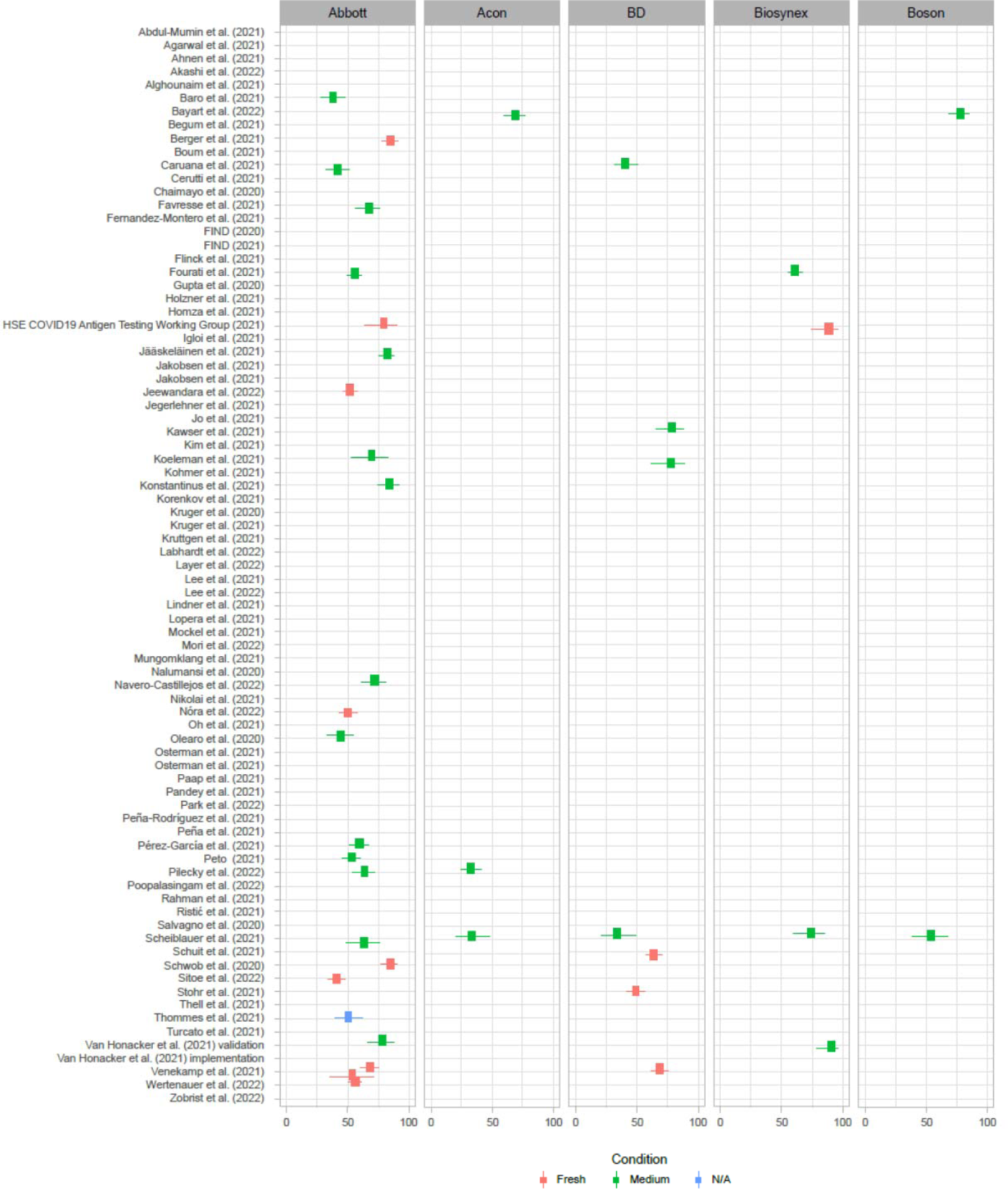
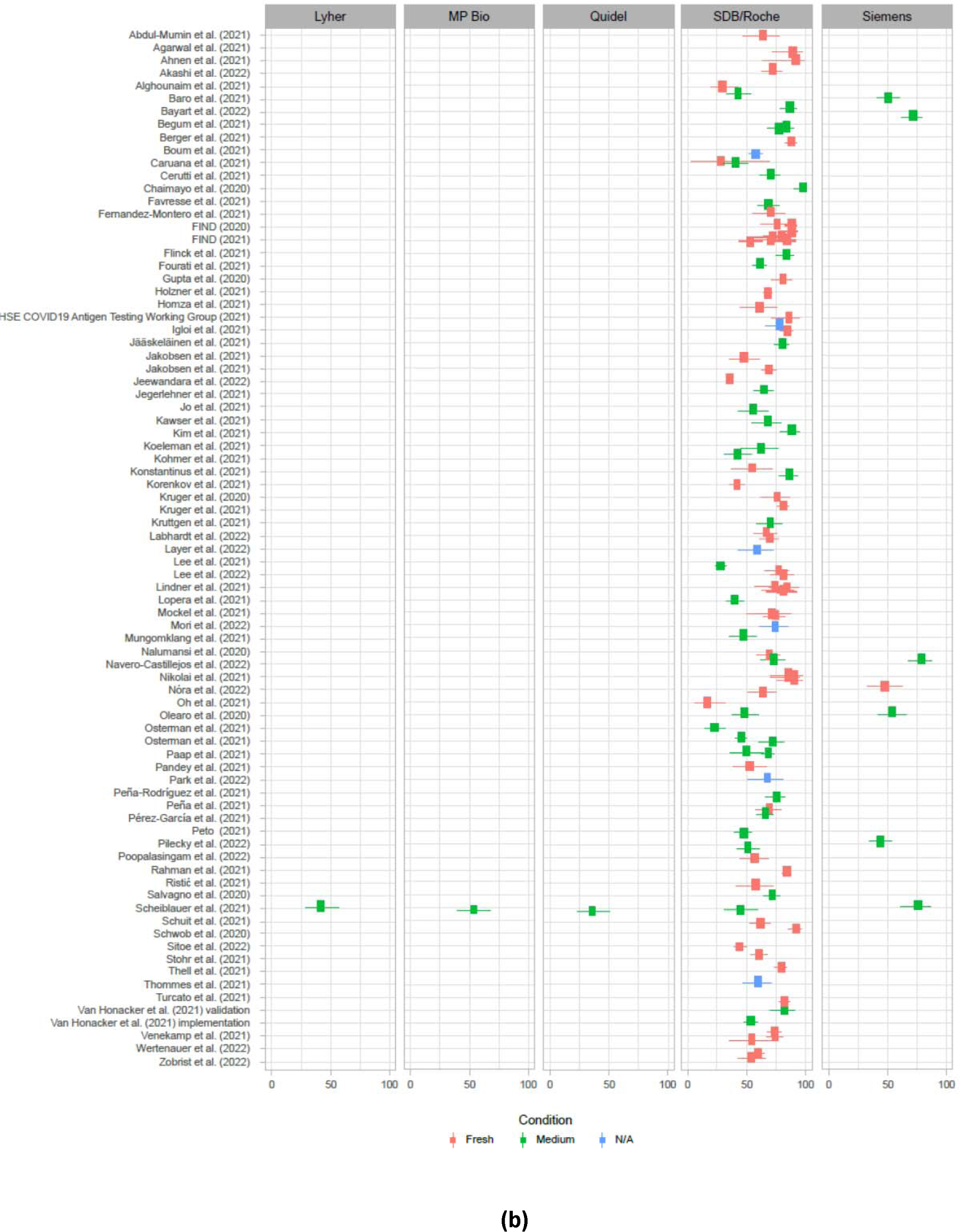
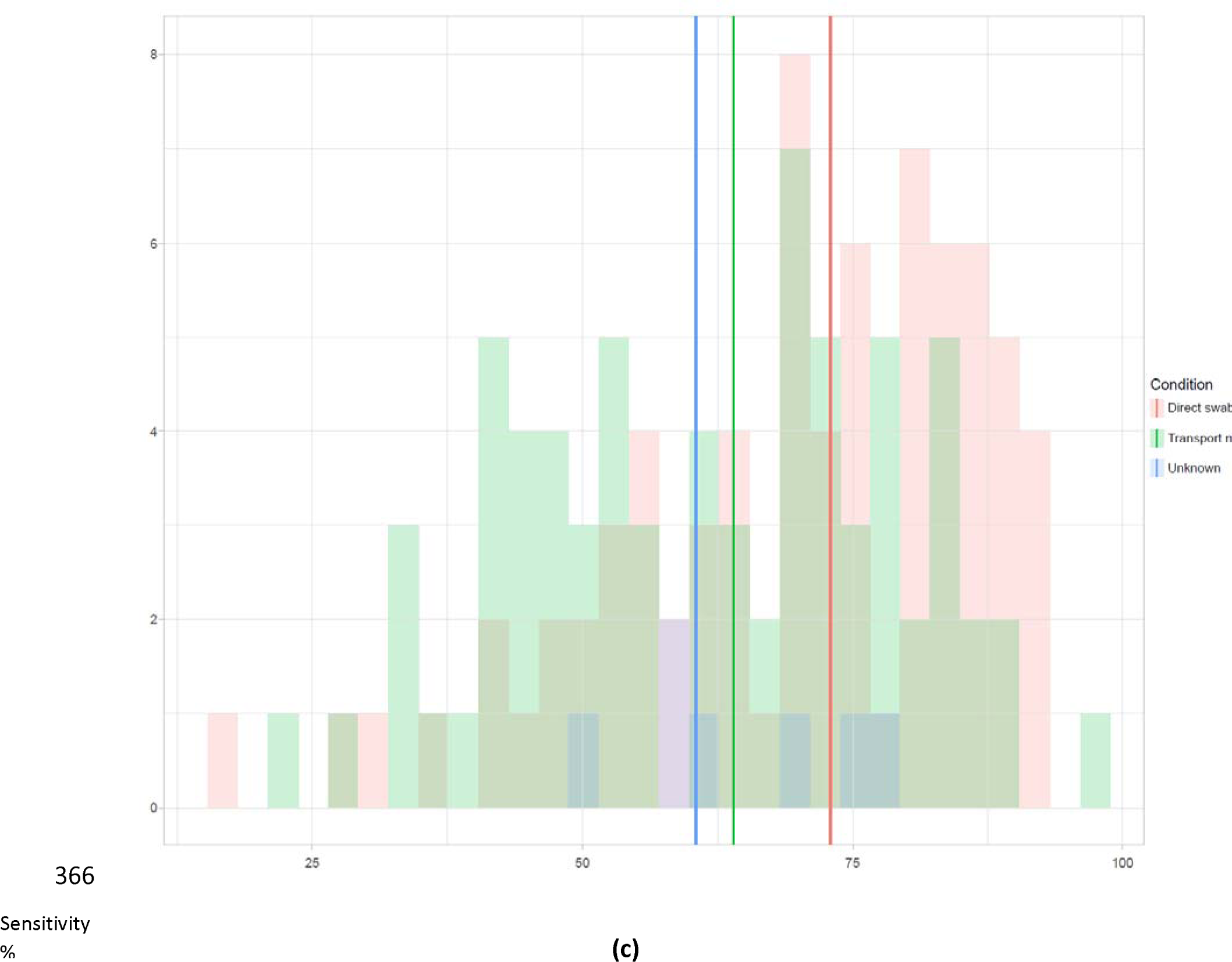
(a) Forest plot of studies showing sensitivity across the cycle (Ct) threshold range grouped by Ct thresholds (≤20, ≤25, ≤30 and ≤33). (b) Forest plot of studies showing sensitivity for direct swabs versus samples stored in transport media, grouped by RAT manufacturer. (c) Histogram of sensitivities for direct swabs versus samples stored in transport media.

A bivariate model was fitted as a linear mixed model, and variance components were estimated by restricted maximum likelihood, using the reitsma function from the “mada” package[16, 17] for each system investigated in five or more studies.[18] The results are presented as a summary receiver operating characteristic (SROC) curve plot **(Figure 4).** The summary estimates, SROC curves, and confidence regions are depicted when a sufficient number of studies was available (>5 studies for SROC curves, >3 studies for summary estimates and confidence regions).

**Figure 4:**
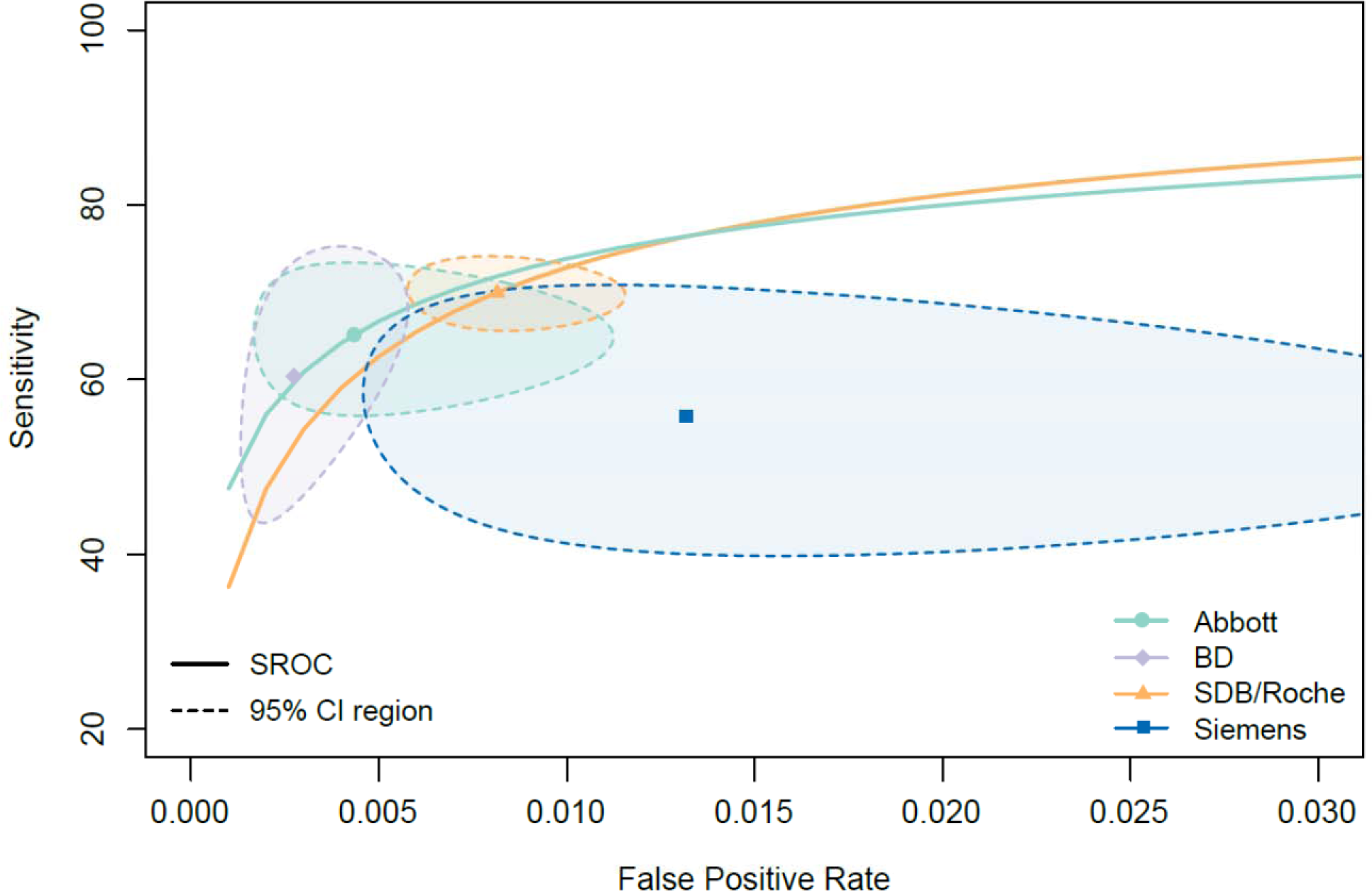
SROC plots for all RATs across different manufacturers. The summary estimate is shown as a filled marker and the associated 95% confidence ellipse as thin dashed lines for each RAT that was covered in more than 3 studies. The SROC curve (thick solid lines) was plotted for all RATs that were covered by more than 5 studies.

## RESULTS

Per PRISMA guidelines, the selection of studies by applying the eligibility criteria is summarized in **Supplemental (Suppl.) Figure 1**. According to our search criteria, 97 publications investigating 166,561 samples were included in this systematic review and qualitative analysis. If the Roche/SDB RAT was compared with one of the 9 common antigen tests of interest, the data from these tests were included as well. The search results included the following antigen tests of interest: i) Panbio**™** COVID-19 Ag Rapid Test Device by Abbott (henceforth called “**Abbott**”); ii) Flowflex**^®^**SARS-CoV-2 Antigen rapid test and COVID-19 Antigen Home Test by Acon Laboratories (henceforth called “**Acon**”); iii) BD Veritor™ System for Rapid Detection of SARS-CoV-2 by Becton Dickinson (henceforth called “**BD**”); iv) BIOSYNEX COVID-19 Ag BSS by Biosynex SA (henceforth called “**Biosynex**”); v) Rapid SARS-CoV-2 Antigen Test Card by Xiamen Boson Biotech (henceforth called “**Boson**”); vi) LYHER^®^ Novel Coronavirus (COVID-19) Antigen Test Kit (Colloidal Gold) by Hangzhou Laihe Biotech (henceforth called “**Laihe**”); vii) Rapid SARS-CoV-2 Antigen Test Card by MP Biomedicals (henceforth called “**MP Bio**”); viii) CLINITEST^®^ Rapid COVID-19 Antigen Test by Siemens Healthineers (henceforth called “**Siemens**”) and ix) Sofia SARS Antigen FIA by Quidel Corporation (henceforth called “**Quidel**”).

The Roche/SDB RAT was investigated in 36 countries within the selected time period. We analyzed the number of studies investigating RAT performance by publication date and geographical location (**Figure 5**). The number of identified studies increased steadily through Q2 2021 as the pandemic progressed. Europe was the leading region evaluating Roche/SDB SARS-CoV-2 RAT performance (n= 65 records), with Asia a distant second (n= 21 records). Studies published in 2021 reported higher numbers of participants compared to 2020, and this is likely due to the increased incidence of SARS-CoV-2 as well as the broader acceptance and availability of RATs (**Figure 5(b)**).

**Figure 5:**
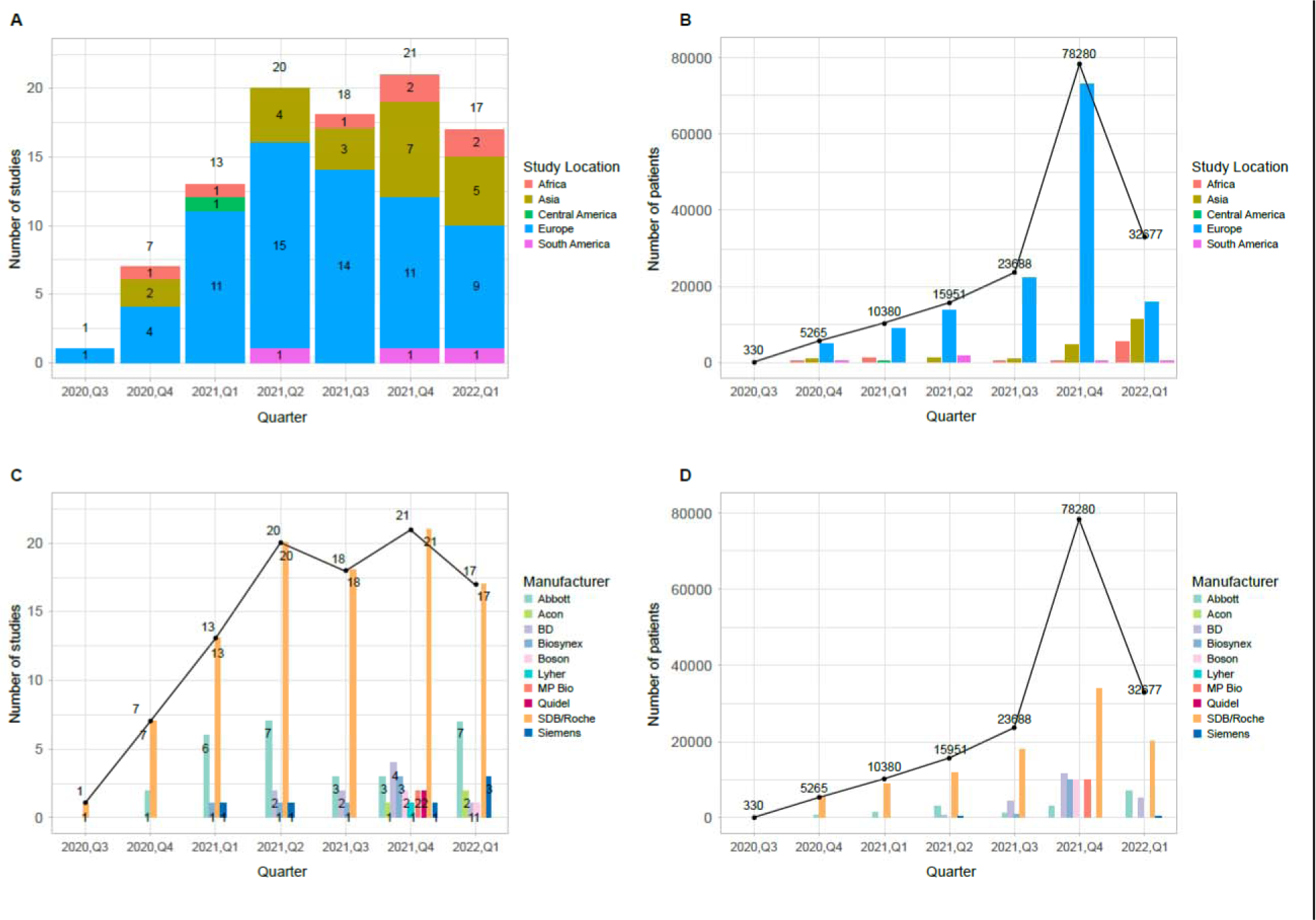
Global evaluation of SARS-CoV-2 rapid antigen tests (RATs) by publication date and geographical location. (a) Distribution of included studies temporally and regionally (b) Distribution of the included study participants temporally and regionally. (c) Distribution of included studies temporally by RAT manufacturer. (d) Distribution of included study participants temporally by RAT manufacturer.

Although the number of relevant studies may have reached a plateau, more participants were recruited in clinical trials in Q1 2022 (n=32,677) compared to Q1 2021 (n=10,380). In addition, the number of participants per quarter showed a trend of recruiting more participants per individual study.

### Overall sensitivity

The quantitative analysis of overall sensitivities was performed for a subset of 84 studies, in which the exact TP and FN numbers were known. All studies included the Roche/SDB RAT (104 distinct cohorts), while Abbott was covered by 23 studies (24 cohorts), BD and Siemens by 7, Biosynex by 4, Acon by 3 studies, Boson was covered by 2 studies, and Laihe, MP Bio, and Quidel by 1 study. For these studies, the pooled sensitivity of RATs varied from 36.0% to 79.4% across manufacturers, with the pooled sensitivity for the Roche/SDB RAT of 70.0% (95% CI: 66.5-73.4) as shown in **Figure 1(a) (also Suppl. Figure 2).** Discrepancies and the wide range of RAT sensitivities may have arisen due to several factors and sub-analyses for these parameters were conducted to assess their impact on test performance.

### Sensitivity in Symptomatic vs Asymptomatic Study Participants

The presence of COVID-19 symptoms (as reported by the studies) was used to perform a stratified sensitivity analysis as shown in **Figure 2(a) (also Suppl. Figure 3)**. Thirty-five studies included symptomatic subjects whereas 26 studies included asymptomatic participants. The median sensitivity of RATs in symptomatic patients was considerably higher (78.0% [95% CI: 73.3-82.7]) than in those without symptoms (49.6% [95% CI: 38.6-58.7]), suggesting that RATs are more effective in detecting an infection in the symptomatic population. This is consistent with previous studies reporting higher RAT sensitivity in the symptomatic population.[19, 20]

The Roche/SDB RAT demonstrated a median sensitivity of 78.9% [95% CI: 74.4-83.4] in symptomatic patients (n=43 distinct cohorts from 35 studies), and a median sensitivity of 53.3% [95% CI: 43.6-63.0] in asymptomatic subjects (n=27 distinct cohorts from 26 studies). Thirteen studies^2^ included RATs from other manufacturers, and reported median sensitivity between 68.6% - 90.4% in symptomatic cohorts and between 27.3% - 57.3% in asymptomatic cohorts.

### Sensitivity for Days Post Symptom Onset (DPSO)

Time elapsed since symptom onset is a clinically relevant parameter.[9, 21–23] Therefore, we analyzed the performance of RATs with regard to DPSO in symptomatic populations. The results showed the following median sensitivity of RATs overall: 85.9% [95% CI: 79.2-92.3], 86.1% [95% CI: 81.2-88.2] and 50.0% [95% CI: 38.2-62.5] for ≤5, ≤7 and >7 DPSO, respectively (**Figure 2(b), also Suppl. Figure 4)**.

The Roche/SDB RAT showed a very similar median sensitivity profile of 86.0% [95% CI: 79.2-92.9], 85.5% [95% CI: 81.2-88.4] and 57.1% [95% CI: 38.2-66.7] for ≤5, ≤7 and >7 DPSO, respectively.

### Sensitivity by Cycle Threshold (Ct)

RAT performance is commonly expressed as relative sensitivity and specificity against RT-PCR. All included studies used Cycle Threshold (Ct) values of the RT-PCR methods as a proxy for viral load in samples, likely due to the lack of an internationally standardized quantitative method at the time these studies were conducted.

Pooled sensitivities grouped by Ct thresholds for each manufacturer as shown in the forest plot in **Figure 3(a)** ranged from 42.5% to 100% **(also Suppl. Figure 5)**. The median sensitivities of the Roche/SDB RAT for Ct values of ≤20, ≤25 and ≤30 were 100.0% [95% CI: 97.2-100], 96.6% [95% CI: 95.2-98.2] and 85.3% [95% CI: 83.3-88.6], respectively. These results showed that RATs are well suited to detect infected individuals with relatively high viral loads, which have been associated with higher risk of virus transmission.[24, 25] Higher Ct values were generally associated with reduced sensitivities.

### Sensitivity for Direct Swabs vs Samples Stored in Transport Media

The majority of SARS-CoV-2 RATs support performance claims for direct swabs from the upper respiratory tract, which must be eluted in the proprietary buffer supplied by the manufacturer. For some RATs, swabs eluted in specific transport media are also validated and claimed. However, due to practical and usability reasons swab samples eluted in a variety of transport media were often used to validate RAT performance in clinical studies and practice.

Our results show slightly inferior median sensitivity for samples eluted/stored in transport media (63.9% [95% CI: 54.0-70.0]) versus direct swabs (72.0% [95% CI: 68.6-76.6]) across manufacturers, although most of the contributing studies were of the Roche/SDB RAT, as shown in **Figure 3(b)**. The median sensitivity of the Roche/SDB RAT for direct swabs and for samples stored in transport media is 74.4% [95% CI: 69.7-80.3] (61 distinct cohorts from 46 studies) and 66.5% [95% CI: 50.9-72.5] (37 distinct cohorts from 33 studies) respectively.

### Overall Specificity

RATs generally have high specificity. This is also reflected in our meta-analysis. The pooled specificity for all manufacturers ranged from 99.1% to 100%, with the pooled specificity for the Roche/SDB RAT of 99.5% [95% CI: 99.3-99.7] as shown in **Figure 1(b) (also Suppl. Figure 6)**. The presence or absence of symptoms had little effect on RAT median specificity (99.8% [95% CI: 99.4-100] or (99.8% [95% CI: 99.6-100]), respectively). Also, the use of transport media did not influence RAT specificity when compared to direct swabs. Our meta-analysis showed equivalent specificity for direct swabs and samples stored in transport media (99.6% and 99.8% respectively) across different manufacturers, although most of the contributing studies were of the Roche/SDB RAT (99.6% [95% CI: 99.3-99.7] and 99.7% [95% CI: 97.7-100] respectively) as depicted in **Suppl. Figure 7.**

### SROC Analyses for all RATs

SROC plots are cumulative ROC plots for data derived from multiple studies that can depict both sensitivity and specificity; for a meaningful analysis a certain sample size is needed. An SROC curve was included only for RATs assessed in more than 5 cohorts, and the overall model estimate per manufacturer with corresponding confidence ellipse only for RATs assessed in more than 3 cohorts. As shown in **Figure 4**, the Roche/SDB, BD and Abbott RATs showed similar performance with largely overlapping CIs while the performance of the Siemens RAT was slightly inferior based on this limited data set (n = 5 studies).

## DISCUSSION

RATs played a critical role during the COVID-19 pandemic. Comprehensive understanding of the capabilities and limitations of RATs for SARS-CoV-2 diagnosis is crucial to assess their utility in an endemic setting. It is important to note that RATs have been shown to be sensitive enough to detect cases with relatively high viral loads, i.e. pre-symptomatic and early symptomatic cases,[26–28] with a slight advantage for direct swabs versus samples stored in transport media.[29] Such cases are likely to account for a significant proportion of transmissions.[24] At present the World Health Organization recommends that RATs meet the minimum performance requirement of at least 80% sensitivity and 97% specificity when compared to NAATs for SARS-CoV-2 detection in symptomatic individuals within the first 7 days of symptoms onset.[23] The US Centers for Disease Control and Prevention (CDC), the European Center for Disease Prevention and Control (eCDC) and the European Union (EU) also set similar requirements for specific cohorts based on symptoms and DPSO, and recommend comparing the RAT results to well characterized and highly sensitive RT-PCR methods.[9, 21, 22]

This global systematic review and meta-analysis presents an overview of the key confounders across studies that report the sensitivity and specificity of commercially available SARS-CoV-2 RATs. Altogether, 97 articles investigating the use of RATs from 10 different manufacturers across 166,561 samples presented findings based on Ct and DPSO ranges, for direct swabs versus samples stored in transport media, and for symptomatic versus asymptomatic patients. The performance of RATs was calculated as relative sensitivity and specificity against RT-PCR results. RATs were typically more sensitive for symptomatic patients, when used less than 7 days after symptom onset, and when using direct swabs. Particularly in samples with a relatively high viral load, which has been associated with increased transmission probability, [24] RATs demonstrated convincing and reliable sensitivity. RAT specificity was generally very high and independent of the parameters assessed in this meta-analysis. The Roche/SDB RAT performed competitively based on the studies assessed, and we found significant differences in test performance across RATs in various studies, though the sample size for some RATs was very small.

We did not aim to assess statistical significance, but the multiple trials selected for systematic review and meta-analysis have a widespread distribution covering a range of prevalence and patient populations, thus the results are generalizable on a larger scale. Overall the results support the applicability of RATs for early case detection.

This study has some limitations. The variation in test sensitivity observed across studies may have been caused by the heterogeneity of study populations, with different disease severity levels and sampling at different time points. The use of different sample types, extraction methods and RT-PCR reference tests may also have contributed to this variation. Further, our meta-analysis focused on studies evaluating the Roche/SDB RAT, and only included a small number of studies that compared the Roche/SDB RAT against a selection of tests offered by other manufacturers. Finally, the meta-analysis contains a limited number of studies that tested the Omicron variant of SARS-CoV-2 due to the timing of this search. Assessment of later studies conducted during the Omicron wave is needed to gain greater insight into RAT diagnostic accuracy in this context, although few novel mutations have been identified in the Omicron nucleocapsid protein (target of most RATs) compared to earlier variants.[7]

In conclusion, the diagnostic accuracy of the SARS-CoV-2 Roche/SDB rapid antigen test in detecting SARS-CoV-2 infections was robust and reliable, especially in samples with a relatively high viral load. Based on 86 studies including comparisons to other RATs, the Roche/SDB RAT showed competitive performance. These results support the use of RATs for early detection of infections and identification of individuals with significant viral loads, thereby contributing to patient management as well as transmission-control decision making.

## Data Availability

All data produced in the present study are available upon reasonable request to the authors

## Acknowledgements

The authors would like to acknowledge the contribution of Sumedha Sinha (Roche Diagnostics Solutions, Pleasanton, California, USA) for writing assistance and proofreading of the article.

## Ethical Approval

The data supporting this manuscript are from published studies, each reporting their own IRB approvals.

## Disclaimer

The Roche/SD Biosensor SARS-CoV-2 Rapid Antigen Test/SARS-CoV-2 Rapid Antigen Test Nasal/ SARS-CoV-2 Antigen Self Test Nasal (Roche/SDB RAT) is an in vitro diagnostic test in countries accepting the CE mark.

## APPENDIX 1 Supplemental Figures

**Supplemental Figure 1:**
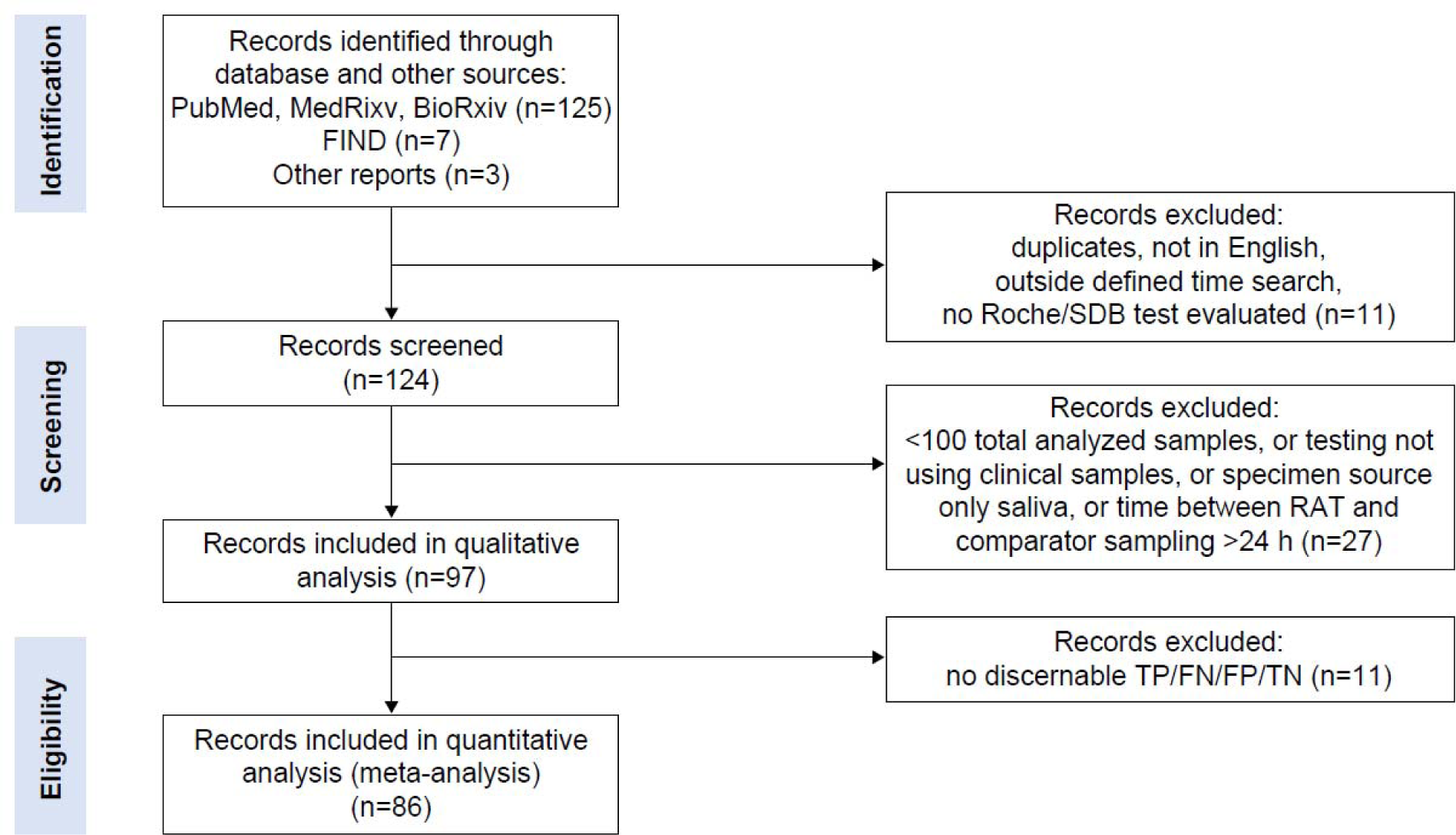
PRISMA Flow chart of study selection for inclusion in the meta-analysis and systematic review.

**Supplemental Figure 2:**
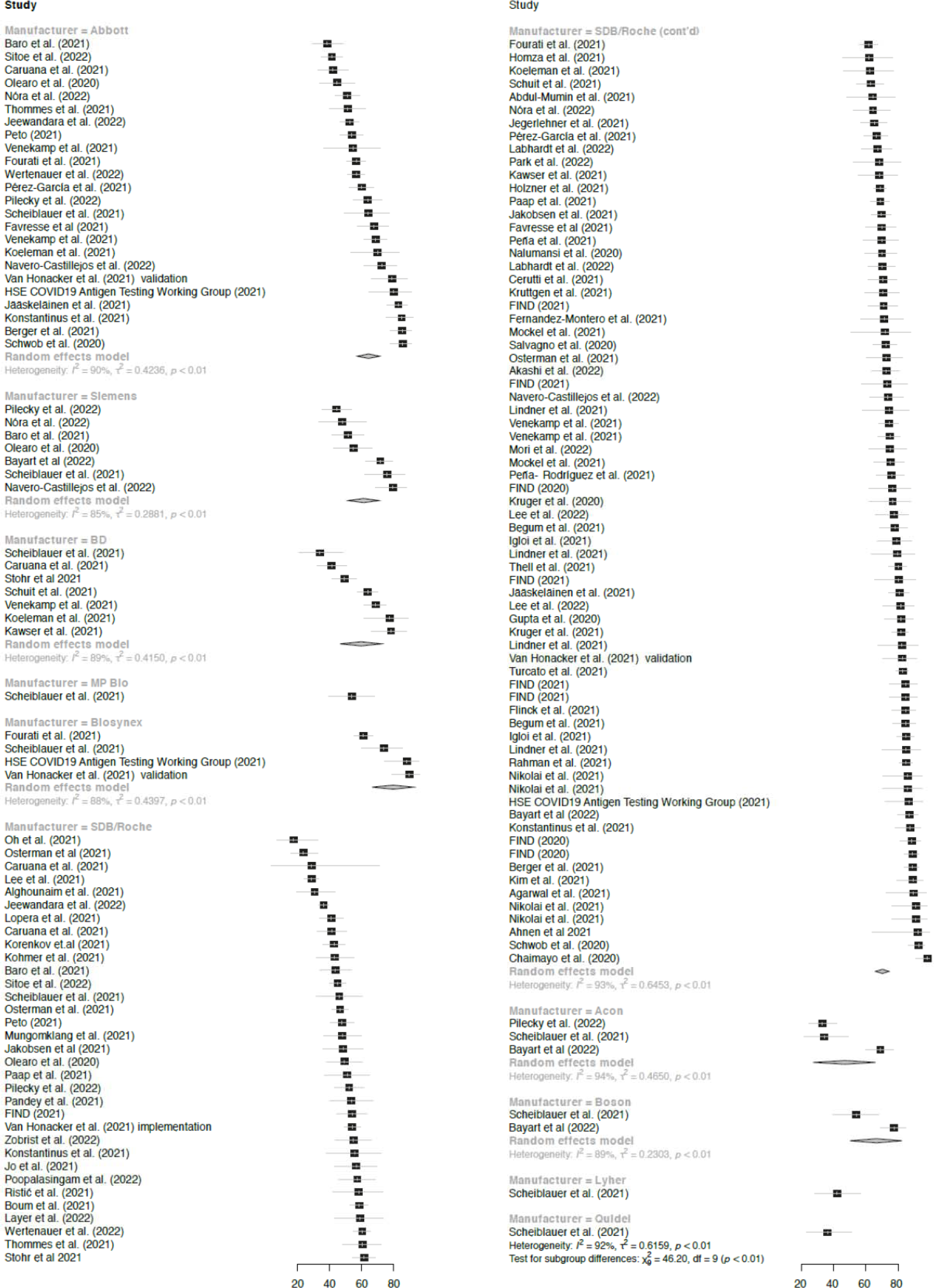
Overall RAT sensitivity across all studies included in the meta-analysis.

**Supplemental Figure 3:**
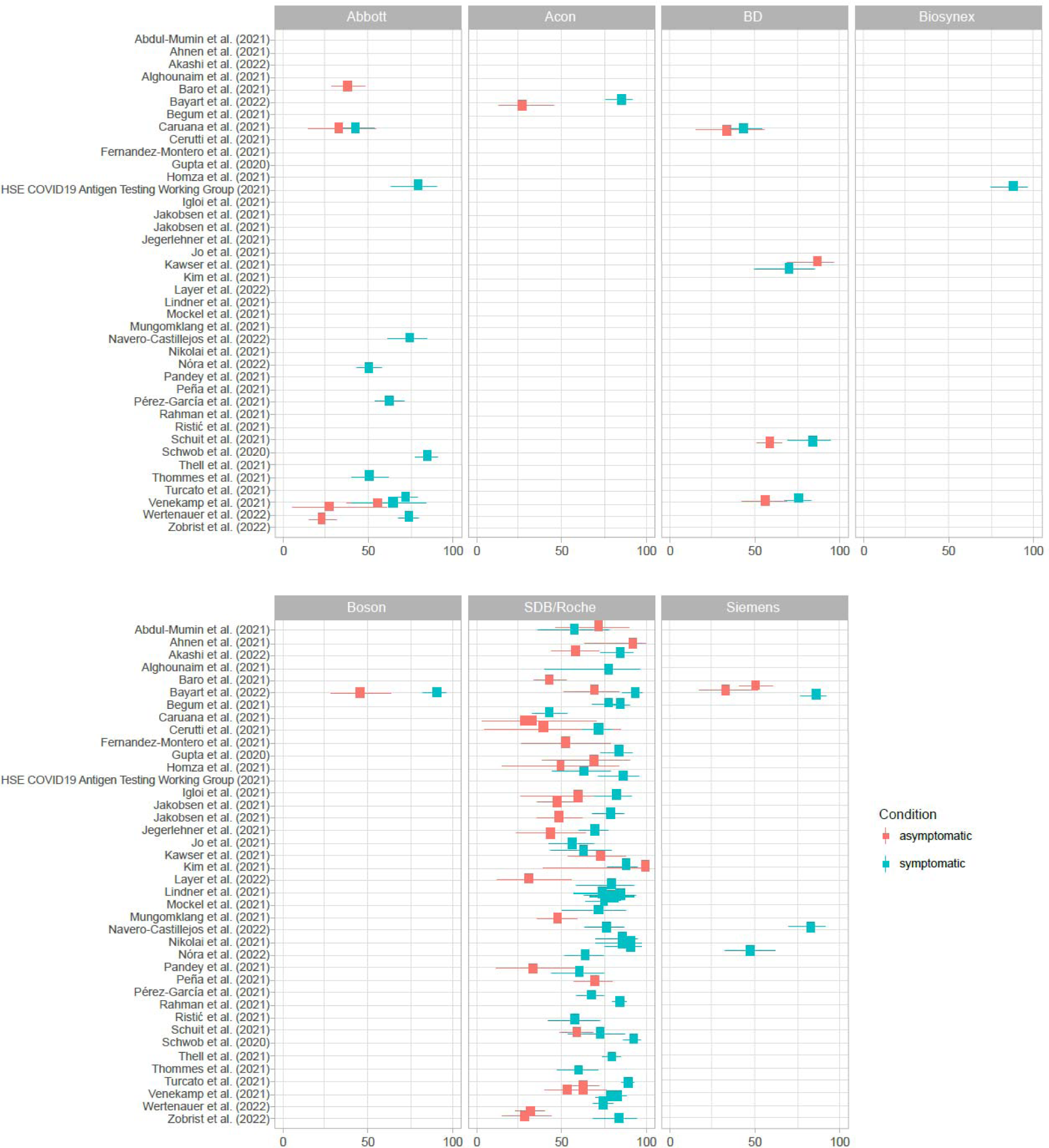
Sensitivity across studies for different RAT manufacturers with symptomatic vs asymptomatic cases coded by color.

**Supplemental Figure 4:**
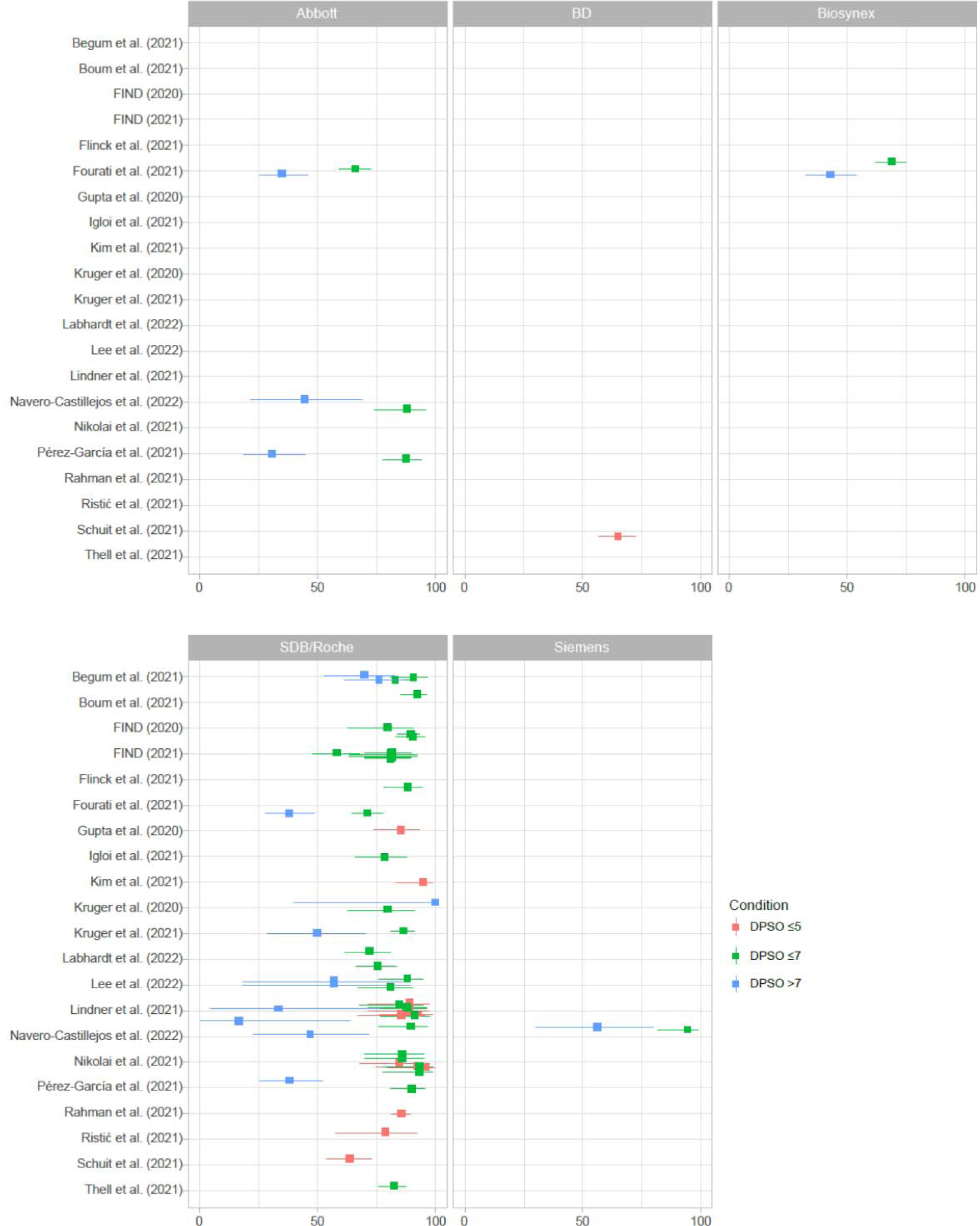
Sensitivity across studies for different RAT manufacturers with number of days post symptom onset (DPSO: ≤5, ≤7, >7) coded by color.

**Supplemental Figure 5:**
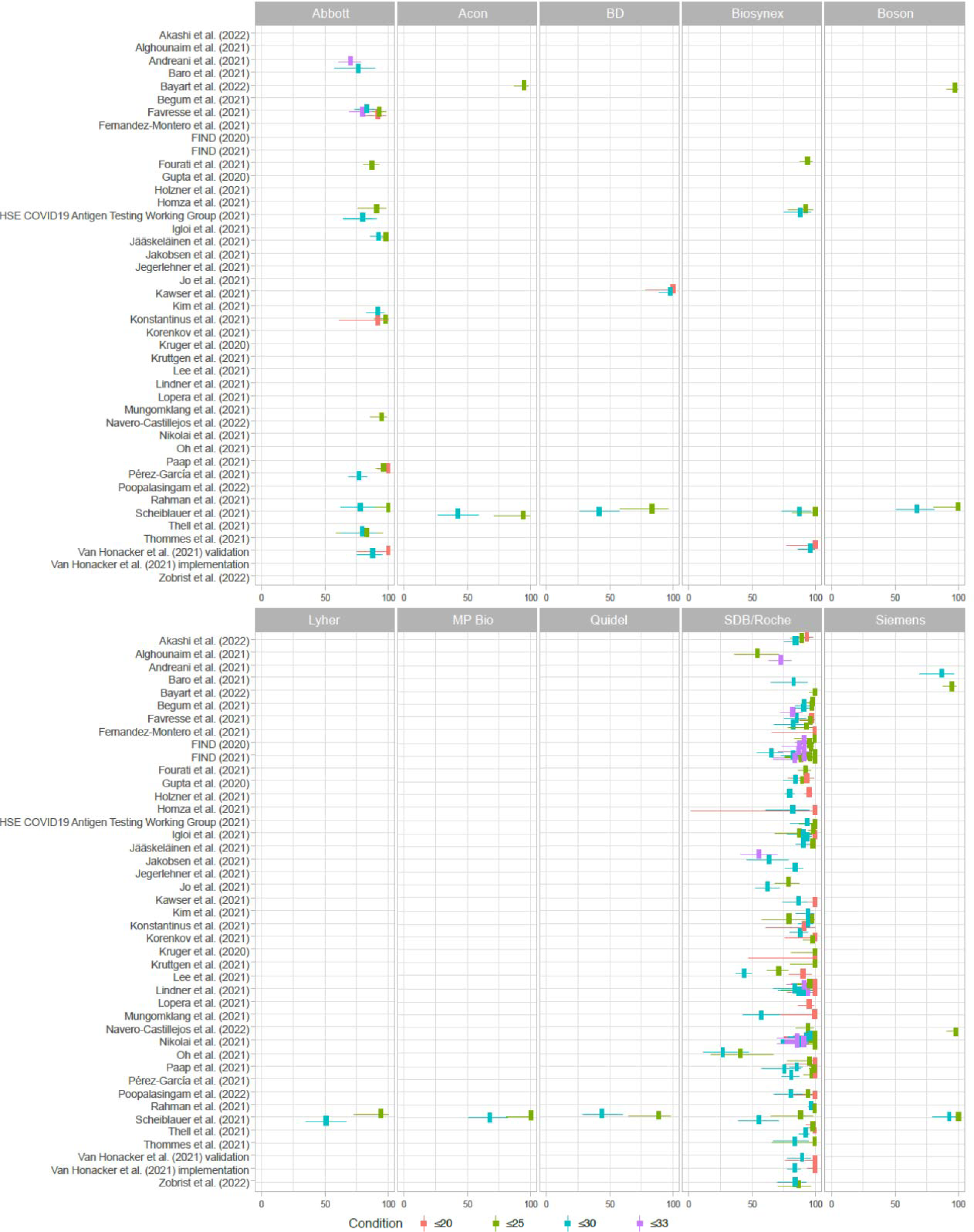
Sensitivity across studies for different RAT manufacturers with cycle threshold (Ct) value (≤20, ≤25, ≤30, ≤33) coded by color.

**Supplemental Figure 6:**
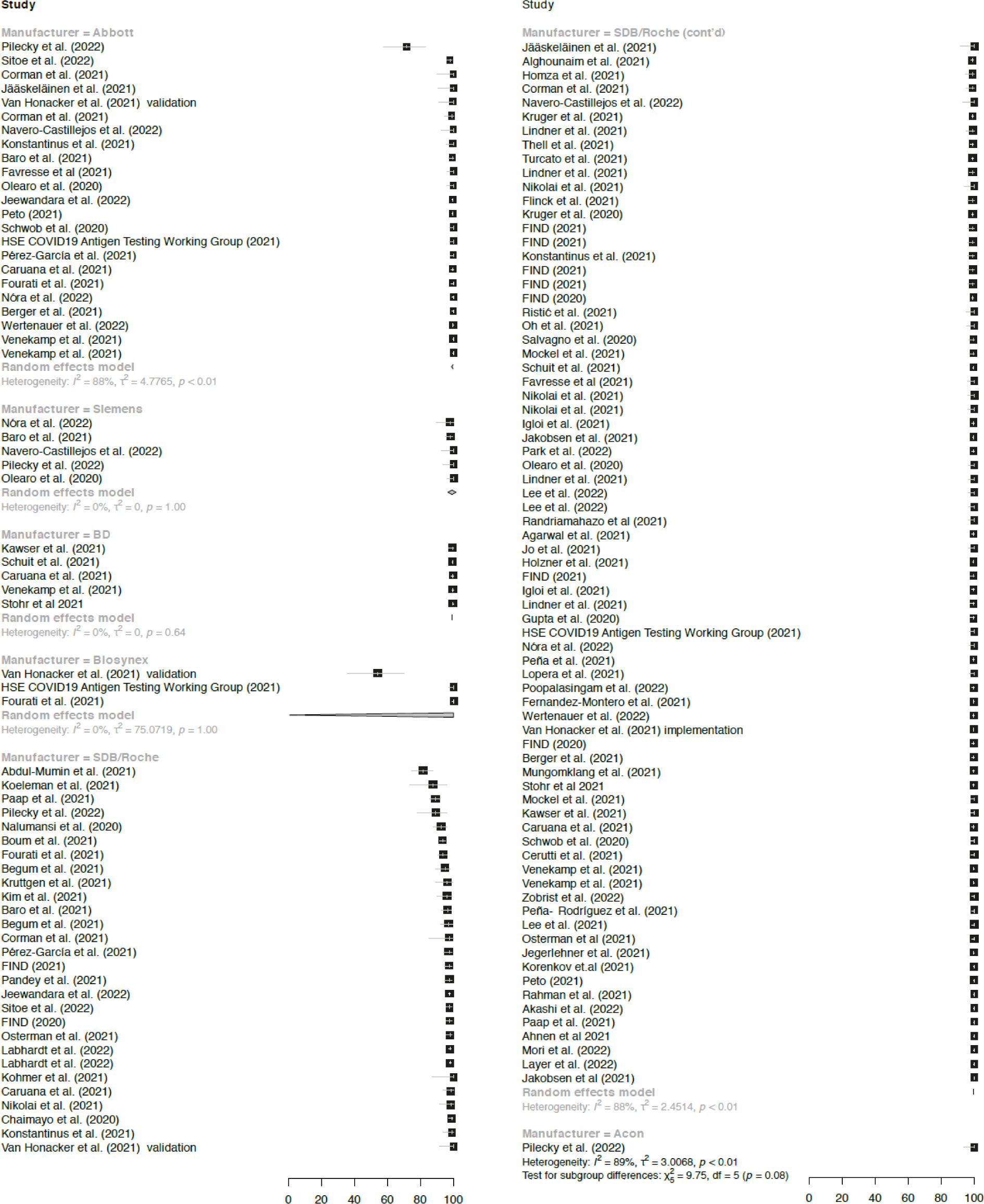
Overall RAT specificity across all studies included in the meta-analysis.

**Supplemental Fig. 7:**
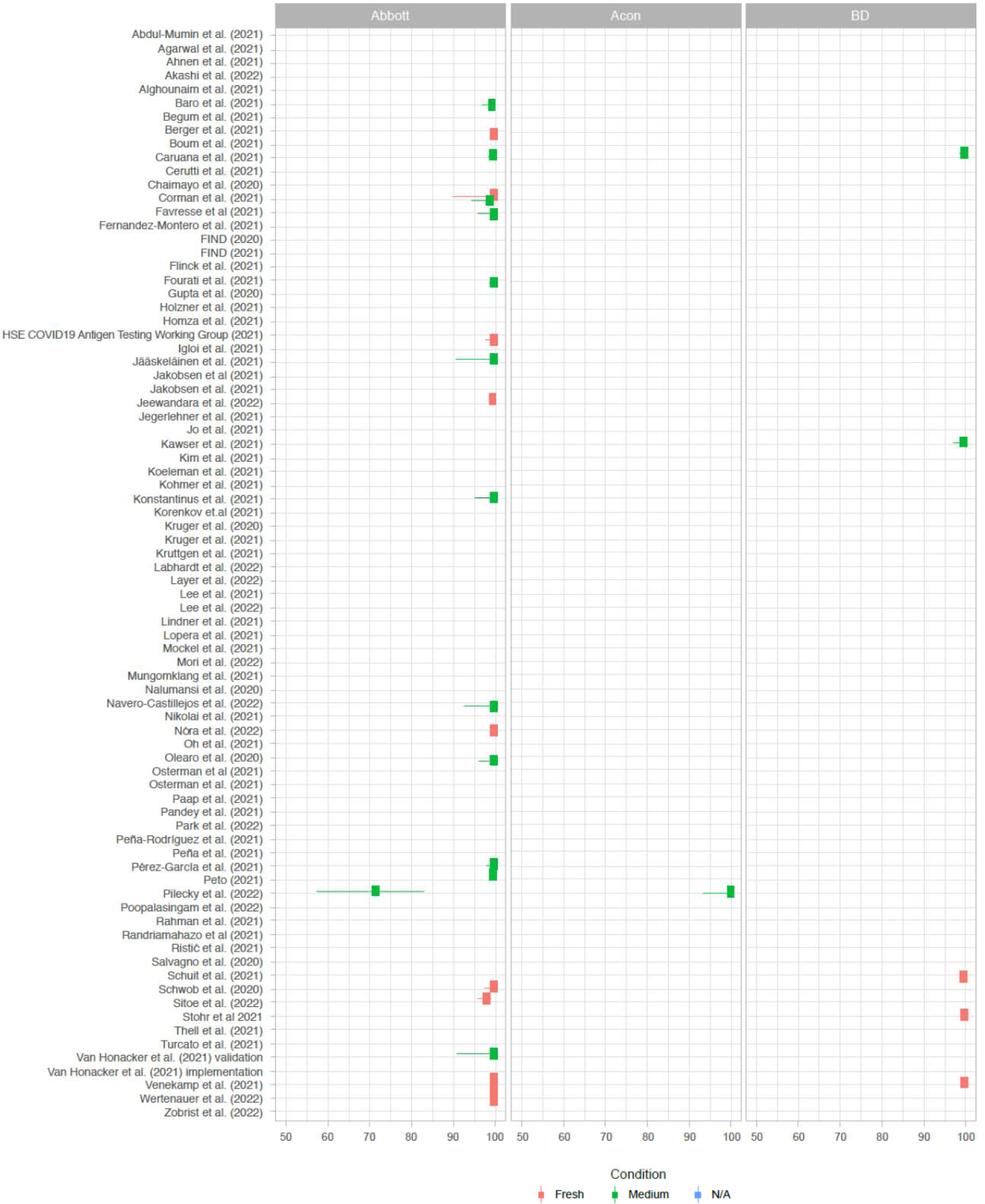
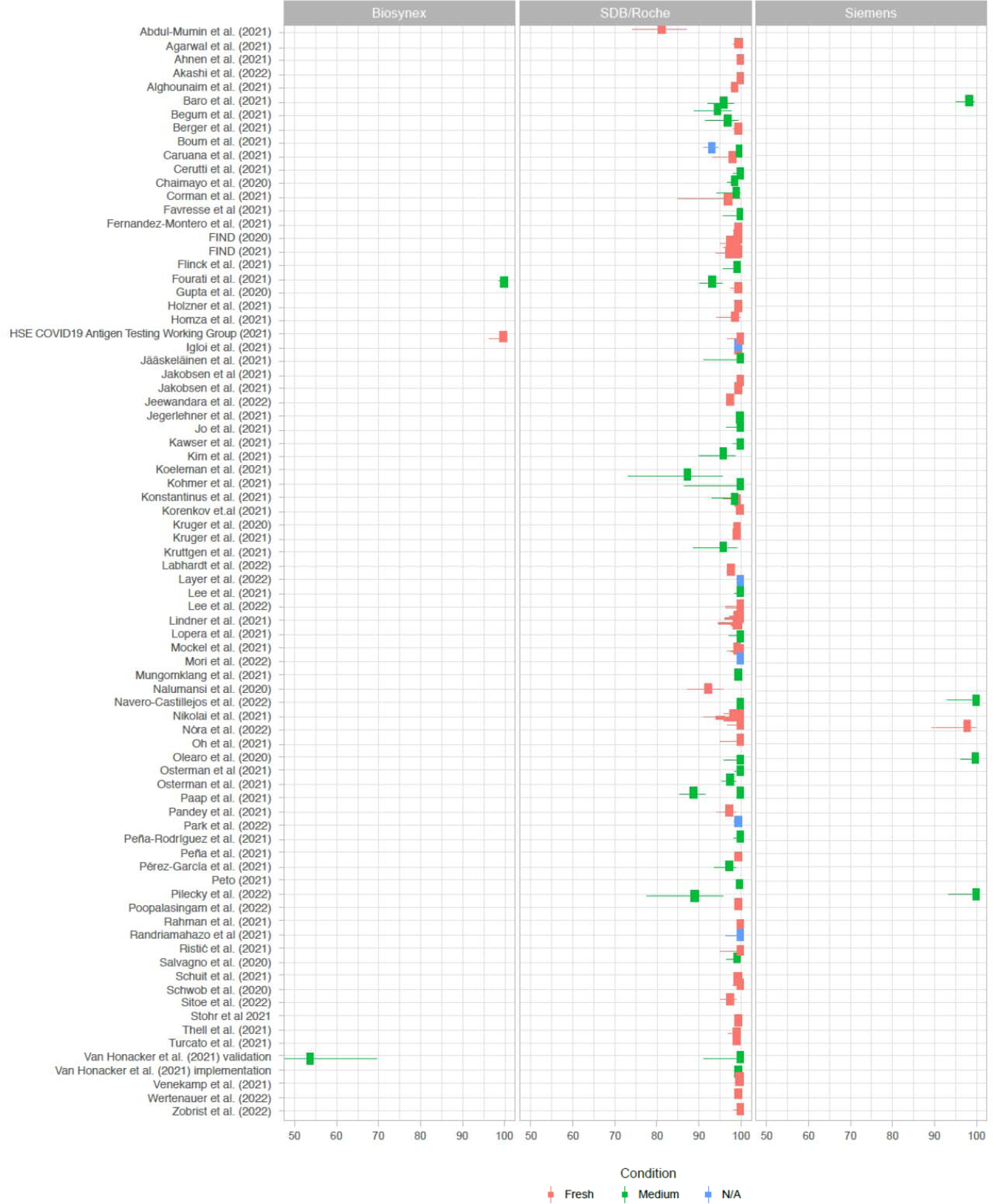
Specificity across studies for different RAT manufacturers with sample type (stored in medium, direct swab or N/A) coded by color.

## Footnotes

1 Also including SARS-CoV-2 Rapid Antigen Test Nasal and SARS-CoV-2 Antigen Self Test Nasal.

2 These 13 studies included the following subsets: 12 studies for sensitivity in symptomatic patients, and 7 studies for sensitivity in asymptomatic patients,.

